# Incidence and Trends in Ischemic Stroke and Mortality in Patients with and Without Type 2 Diabetes in Taiwan 2000-2018

**DOI:** 10.1101/2023.12.21.23300417

**Authors:** Fu-Shun Yen, James Cheng-Chung Wei, Tzu-Ju Hsu, Ying-Hsiu Shih, Yun Kai Yeh, Chih-Cheng Hsu, Chii-Min Hwu

## Abstract

**BACKGROUND:** Stroke has a significant impact on life expectancy, comparable to that of cardiovascular disease in Asia. We conducted this longitudinal study to examine the incidence of ischemic stroke and 30-day mortality among persons with and without type 2 diabetes (T2D). We also compared outcomes between those over and under 60 years and between men and women.

**METHODS:** We enrolled participants from the 2000 to 2018 National Health Insurance Research Database in Taiwan. Cox proportional-hazard models were used to examine the outcomes.

**RESULTS:** The incidence of in-hospital ischemic stroke in patients with and without T2D had significant reductions of 64.42% and 55.63%, respectively. The relative risk of stroke associated with diabetes remained significantly higher (2.01 to 2.33 times) than non-diabetes. The relative risk of stroke in patients under 60 with T2D was 2.56 to 4.36 times higher than in those without T2D. Between 2000 and 2018, there was a significant 83.24% and 88.55% reduction in the risk of 30-day mortality after stroke in patients with and without T2D. There was no significant increase in the risk of 30-day mortality in people with diabetes compared to those without diabetes. However, the relative risk of death from stroke in patients under 60 years with diabetes was 1.63 to 2.49 times higher than in those without diabetes.

**CONCLUSIONS:** This nationwide cohort study showed that the incidence of in-hospital ischemic stroke and 30-day mortality in patients with and without T2D exhibited a significant decreasing trend from 2000 to 2018. Patients with T2D had about twice the relative risk of ischemic stroke compared to those without T2D.

## INTRODUCTION

According to the Global Burden of Disease Study, stroke is the third leading cause of long-term disability and mortality worldwide.^1^ There are 101.5 million cases of stroke worldwide, resulting in approximately 6.6 million cases of premature mortality with 125.4 million years of life lost and 17.7 million years of living with disability in the Global Burden of Disease Study 2019.^2^ Therefore, stroke poses a severe threat of disability and death for adults. Furthermore, stroke risk varies between Asian countries and Western nations, with a higher risk to life and potential for death than cardiovascular disease.^1,3,4^ Type 2 diabetes (T2D) is characterized by hyperglycemia and insulin resistance and is often associated with obesity, hypertension, and dyslipidemia, which increases patient susceptibility to cerebral atherosclerosis and stroke.^5^ According to a meta-analysis and systemic review of 102 prospective studies, people with T2D have a 2.27 times higher risk of ischemic stroke than those without T2D.^6^ Hyperglycemia can lead to impaired recanalization and decreased reperfusion in stroke patients, which results in further brain tissue damage and contributes to a worse prognosis for these patients.^5^

In recent years, some countries have advocated for proactive treatment of diabetes, hypertension, and hyperlipemia, leading to a notable decrease in the incidence and mortality rates of stroke in some countries.^7,8^ However, the incidence of stroke remains unchanged in some countries, while in others, it has increased.^9,10^ Furthermore, individuals with diabetes still have a significantly higher risk of stroke and stroke-related death than those without diabetes.^6,11^ Moreover, a growing trend of young stroke patients (under 60 years) is noted worldwide.^2,5,12^ As in Asian countries, stroke has a significant impact on life, comparable to that of cardiovascular disease.^3^ However, most research on the relationship between diabetes and stroke has been conducted in Western countries, with relatively fewer studies conducted in Eastern countries. Furthermore, many studies do not delve into detailed data on diabetes and stroke in younger individuals and recent mortality figures related to stroke. Therefore, we conducted this longitudinal study to investigate the incidence of ischemic stroke and within 30-day mortality rate among persons with and without T2D in Taiwan from 2000 to 2018.

We also compared the stroke incidence and mortality risks between those over and under 60 and between males and females.

## METHODS

### Data Sources

The government of Taiwan initiated the National Health Insurance (NHI) program in 1995. This mandatory insurance program is largely funded by the government and employers, with only a minimal contribution required from individuals. By 2000, nearly 99% of Taiwan’s 29 million people were enrolled in this insurance initiative. The National Health Insurance Research Database (NHIRD) maintains comprehensive medical records since 1995, documenting details such as patient age, date of birth, sex, place of residence, treatments received, and diagnostic data according to the International Classification of Diseases, Ninth Revision, Clinical Modification (ICD-9-CM) and ICD-10-CM codes. For this study, we methodically recruited individuals from the NHIRD.^13^ The research was approved by the Research Ethics Committee of China Medical University and Hospital [CMUH111-REC2-109(CR-1)]. In accordance with privacy principles, all identifiable information from patients or care providers was encrypted before release, exempting this study from the requirement to obtain informed consent from participants by the Research Ethics Committee.

### Study Design and Population

This nationwide cohort study consecutively enrolled participants from the 2000-2004, 2005-2009, 2010-2014, and 2015-2018 NHIRDs to assess trends in ischemic stroke incidence among individuals with and without T2D. Identification of T2D was based on ICD-9/10-CM codes (Table S1), requiring at least 2 outpatient claims within 1 year or a single hospital admission record. The use of ICD codes for defining T2D is validated by previous studies, with an accuracy rate of 93.3%.^14^

We categorized individuals diagnosed with T2D as study cases, with the day of T2D diagnosis as the index date; those without a T2D diagnosis were control cases, with the same calendar year and corresponding index date for the study subject as the index date for the control cases. Follow-up began on the respective index dates and ended on December 31, 2019.

We excluded individuals identified with type 1 diabetes, those younger than 20 years or older than 100 years, those with missing data on age or sex, and those who had experienced a stroke or had been diagnosed with gestational diabetes mellitus before the index date.

### Comorbidities

This study assessed several potential confounding factors, including age, sex, obesity (a composite of overweight, obese, or severely obese), smoking habits, alcohol-related disorders, hypertension, dyslipidemia, coronary artery disease, heart failure, valvular heart disease, atrial fibrillation, chronic kidney disease, chronic obstructive pulmonary disease (COPD), liver cirrhosis, peripheral artery disease, history of venous thromboembolism, connective tissue disease, hyperthyroidism, and neoplasms. We also included the Charlson Comorbidity Index (CCI).^15^ and Diabetes Complication Severity Index (DCSI)^16^ scores to assess participants’ disease burden. We considered medications such as insulin, type and amount of antidiabetic and antihypertensive agents, statin, aspirin, ADP receptor inhibitors, vitamin K antagonists, and direct oral anticoagulants (Table 1).

**Table 1.**
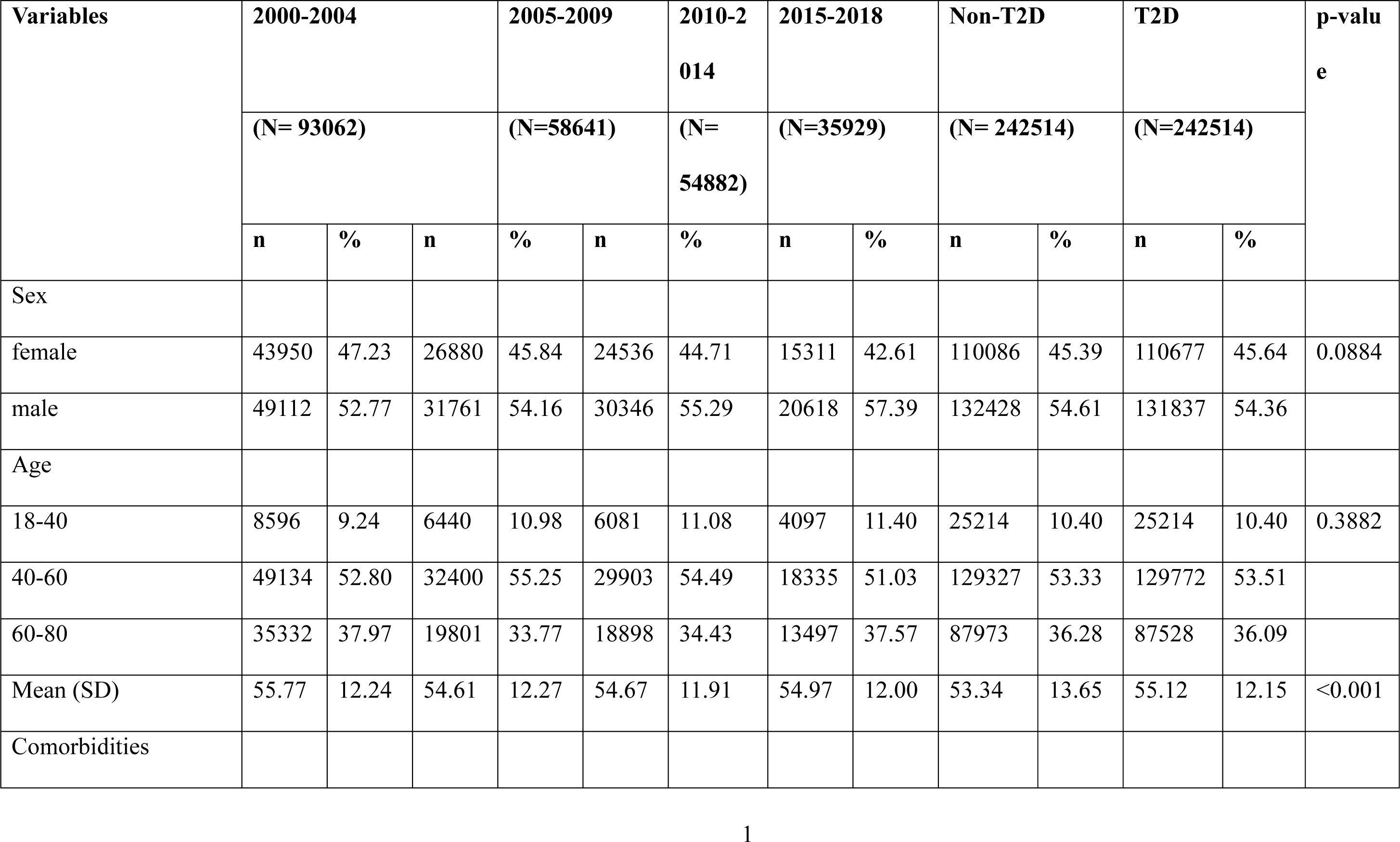
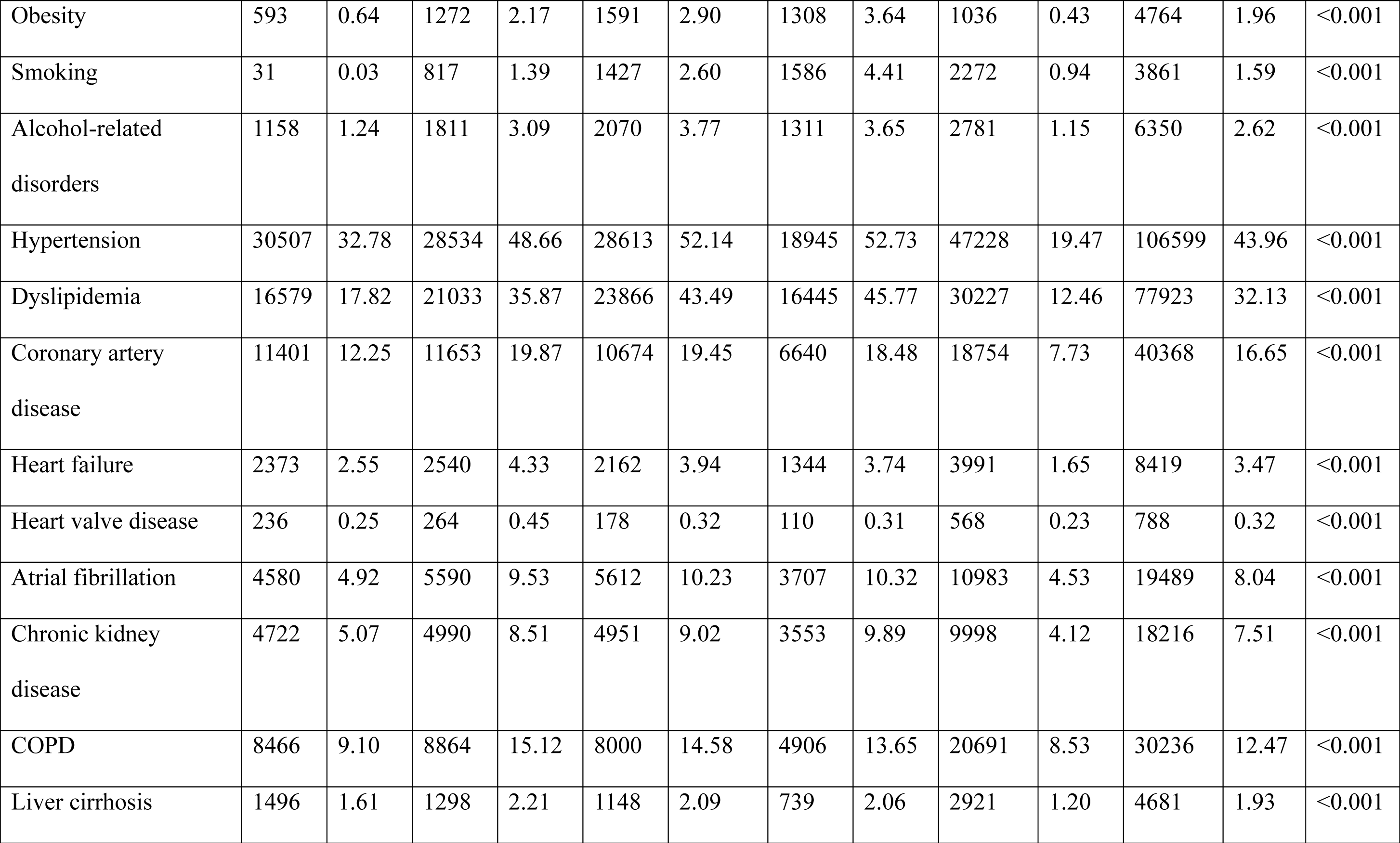
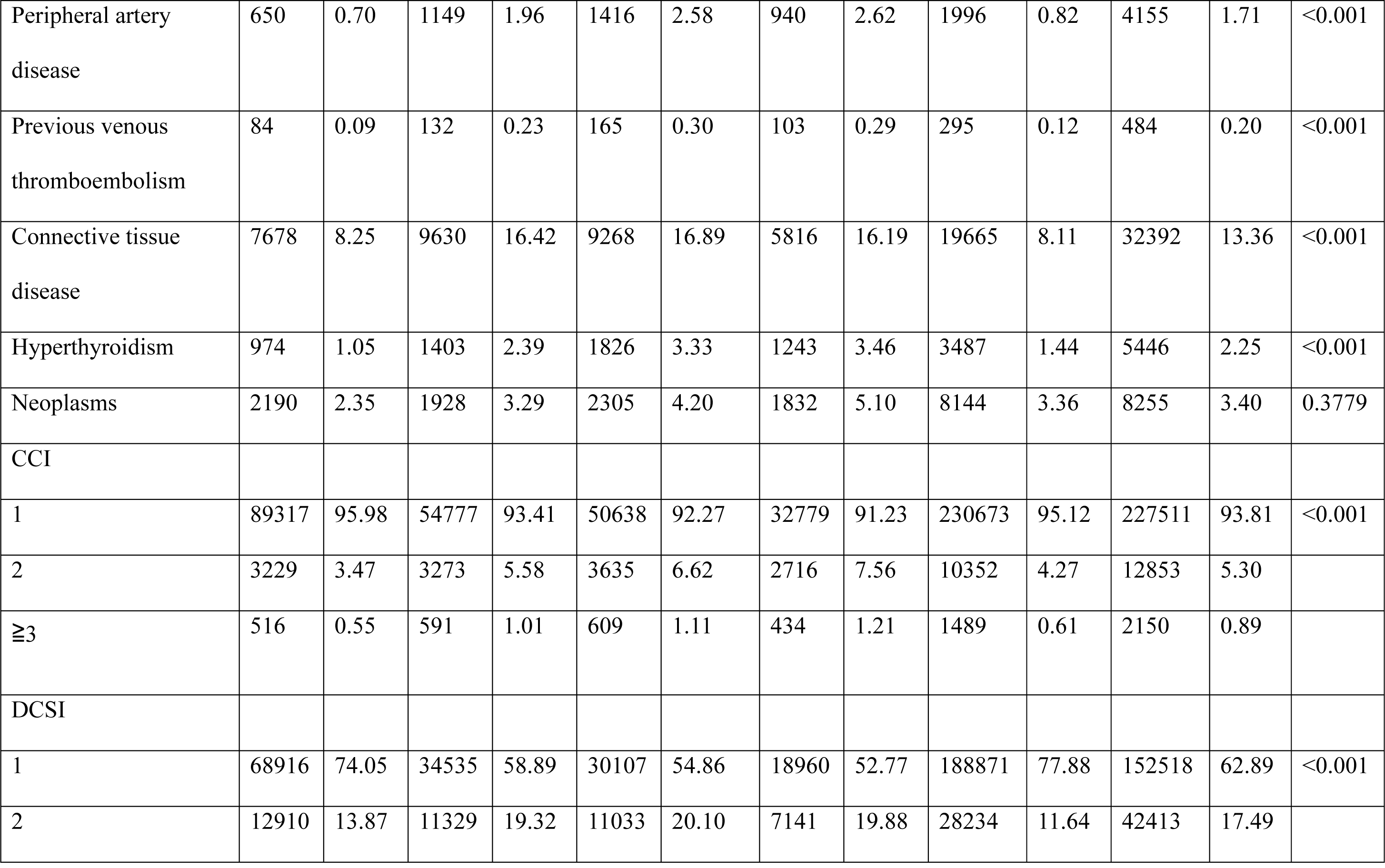
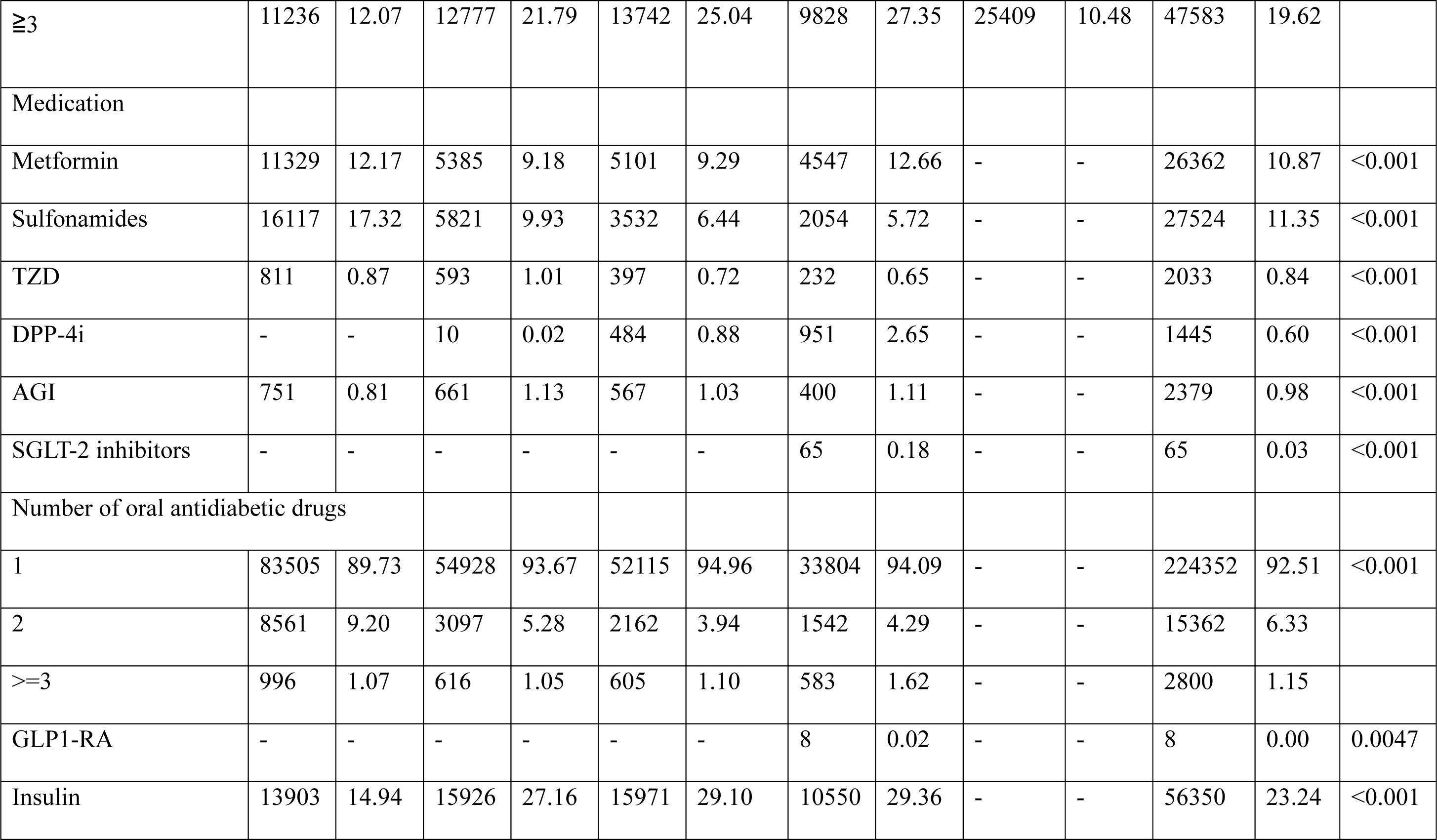
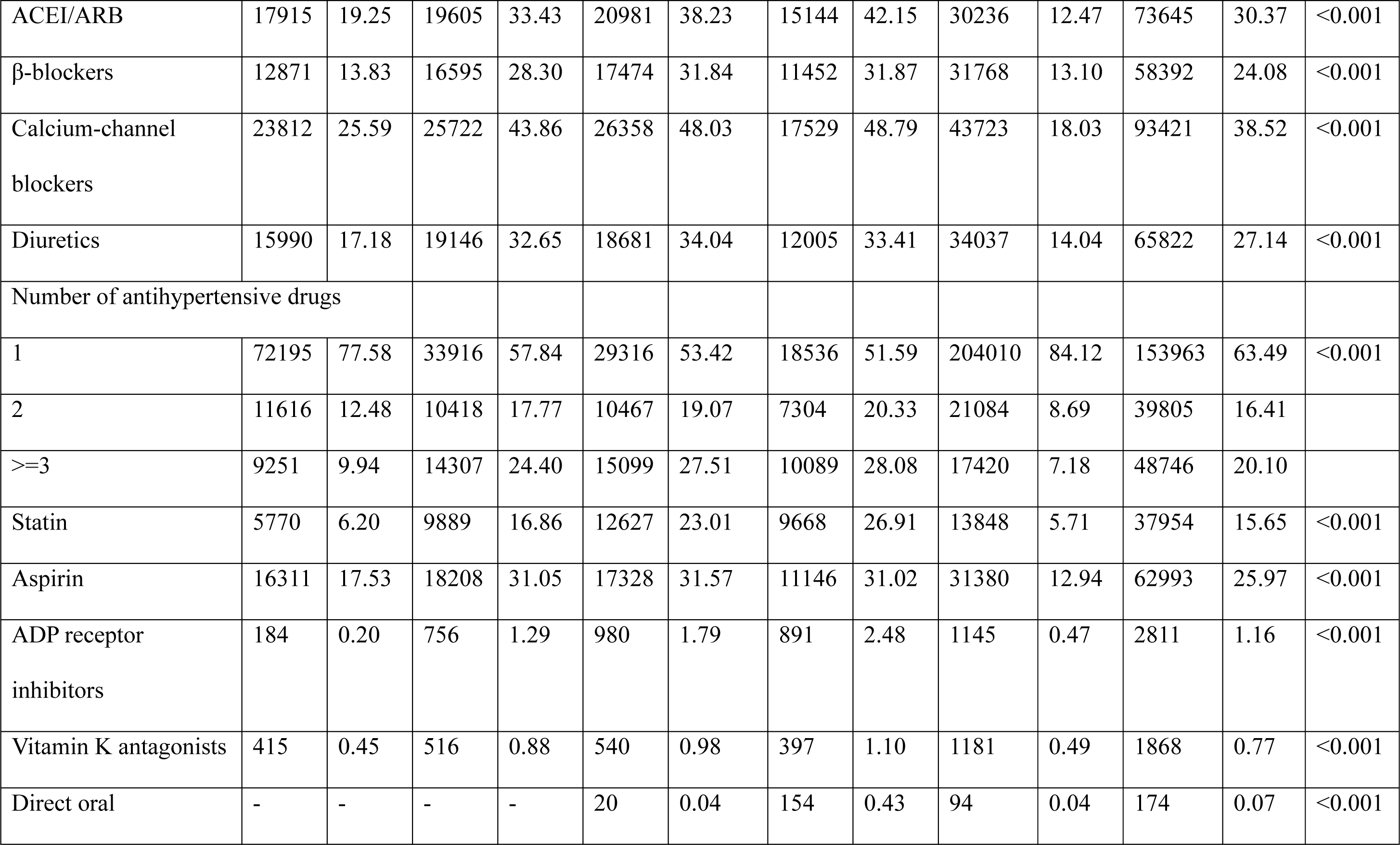
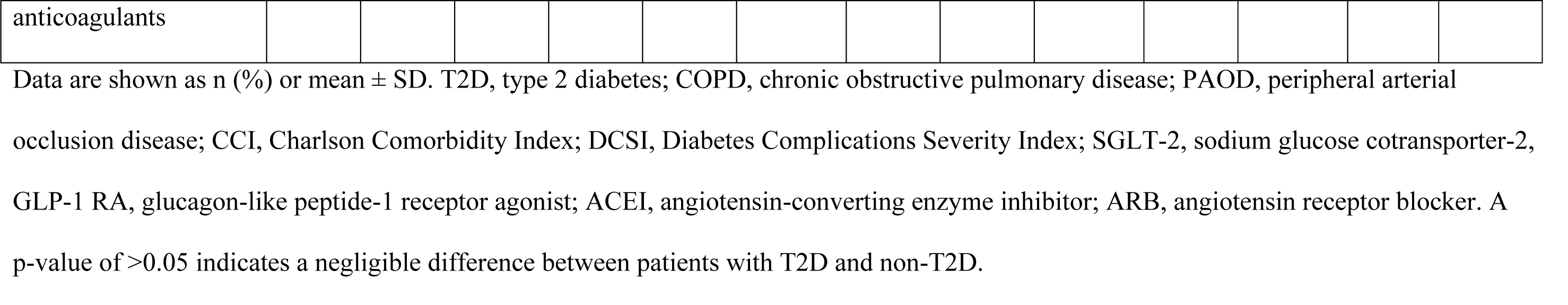
Baseline Characteristics and Comorbidities in Persons with and Without T2D in Taiwan from 2000 to 2018.

| Variables | 2000-2004 |  |  | 2005-2009 |  | 2010-2014 | 2015-2018 |  | Non-T2D |  | T2D |  | p-value |
| --- | --- | --- | --- | --- | --- | --- | --- | --- | --- | --- | --- | --- | --- |
|  | (N= 93062) |  |  | (N=58641) |  | (N=54882) | (N=35929) |  | (N= 242514) |  | (N=242514) |  |  |
|  | n | % | n | % | n | % | n | % | n | % | n | % |  |
| Sex |  |  |  |  |  |  |  |  |  |  |  |  |  |
| female | 43950 | 47.23 | 26880 | 45.84 | 24536 | 44.71 | 15311 | 42.61 | 110086 | 45.39 | 110677 | 45.64 | 0.0884 |
| male | 49112 | 52.77 | 31761 | 54.16 | 30346 | 55.29 | 20618 | 57.39 | 132428 | 54.61 | 131837 | 54.36 |  |
| Age |  |  |  |  |  |  |  |  |  |  |  |  |  |
| 18-40 | 8596 | 9.24 | 6440 | 10.98 | 6081 | 11.08 | 4097 | 11.40 | 25214 | 10.40 | 25214 | 10.40 | 0.3882 |
| 40-60 | 49134 | 52.80 | 32400 | 55.25 | 29903 | 54.49 | 18335 | 51.03 | 129327 | 53.33 | 129772 | 53.51 |  |
| 60-80 | 35332 | 37.97 | 19801 | 33.77 | 18898 | 34.43 | 13497 | 37.57 | 87973 | 36.28 | 87528 | 36.09 |  |
| Mean (SD) | 55.77 | 12.24 | 54.61 | 12.27 | 54.67 | 11.91 | 54.97 | 12.00 | 53.34 | 13.65 | 55.12 | 12.15 | <0.001 |
| Comorbidities |  |  |  |  |  |  |  |  |  |  |  |  |  |
| Obesity | 593 | 0.64 | 1272 | 2.17 | 1591 | 2.90 | 1308 | 3.64 | 1036 | 0.43 | 4764 | 1.96 | <0.001 |
| Smoking | 31 | 0.03 | 817 | 1.39 | 1427 | 2.60 | 1586 | 4.41 | 2272 | 0.94 | 3861 | 1.59 | <0.001 |
| Alcohol-related disorders | 1158 | 1.24 | 1811 | 3.09 | 2070 | 3.77 | 1311 | 3.65 | 2781 | 1.15 | 6350 | 2.62 | <0.001 |
| Hypertension | 30507 | 32.78 | 28534 | 48.66 | 28613 | 52.14 | 18945 | 52.73 | 47228 | 19.47 | 106599 | 43.96 | <0.001 |
| Dyslipidemia | 16579 | 17.82 | 21033 | 35.87 | 23866 | 43.49 | 16445 | 45.77 | 30227 | 12.46 | 77923 | 32.13 | <0.001 |
| Coronary artery disease | 11401 | 12.25 | 11653 | 19.87 | 10674 | 19.45 | 6640 | 18.48 | 18754 | 7.73 | 40368 | 16.65 | <0.001 |
| Heart failure | 2373 | 2.55 | 2540 | 4.33 | 2162 | 3.94 | 1344 | 3.74 | 3991 | 1.65 | 8419 | 3.47 | <0.001 |
| Heart valve disease | 236 | 0.25 | 264 | 0.45 | 178 | 0.32 | 110 | 0.31 | 568 | 0.23 | 788 | 0.32 | <0.001 |
| Atrial fibrillation | 4580 | 4.92 | 5590 | 9.53 | 5612 | 10.23 | 3707 | 10.32 | 10983 | 4.53 | 19489 | 8.04 | <0.001 |
| Chronic kidney disease | 4722 | 5.07 | 4990 | 8.51 | 4951 | 9.02 | 3553 | 9.89 | 9998 | 4.12 | 18216 | 7.51 | <0.001 |
| COPD | 8466 | 9.10 | 8864 | 15.12 | 8000 | 14.58 | 4906 | 13.65 | 20691 | 8.53 | 30236 | 12.47 | <0.001 |
| Liver cirrhosis | 1496 | 1.61 | 1298 | 2.21 | 1148 | 2.09 | 739 | 2.06 | 2921 | 1.20 | 4681 | 1.93 | <0.001 |
| Peripheral artery disease | 650 | 0.70 | 1149 | 1.96 | 1416 | 2.58 | 940 | 2.62 | 1996 | 0.82 | 4155 | 1.71 | <0.001 |
| Previous venous thromboembolism | 84 | 0.09 | 132 | 0.23 | 165 | 0.30 | 103 | 0.29 | 295 | 0.12 | 484 | 0.20 | <0.001 |
| Connective tissue disease | 7678 | 8.25 | 9630 | 16.42 | 9268 | 16.89 | 5816 | 16.19 | 19665 | 8.11 | 32392 | 13.36 | <0.001 |
| Hyperthyroidism | 974 | 1.05 | 1403 | 2.39 | 1826 | 3.33 | 1243 | 3.46 | 3487 | 1.44 | 5446 | 2.25 | <0.001 |
| Neoplasms | 2190 | 2.35 | 1928 | 3.29 | 2305 | 4.20 | 1832 | 5.10 | 8144 | 3.36 | 8255 | 3.40 | 0.3779 |
| CCI |  |  |  |  |  |  |  |  |  |  |  |  |  |
| 1 | 89317 | 95.98 | 54777 | 93.41 | 50638 | 92.27 | 32779 | 91.23 | 230673 | 95.12 | 227511 | 93.81 | <0.001 |
| 2 | 3229 | 3.47 | 3273 | 5.58 | 3635 | 6.62 | 2716 | 7.56 | 10352 | 4.27 | 12853 | 5.30 |  |
| ≥3 | 516 | 0.55 | 591 | 1.01 | 609 | 1.11 | 434 | 1.21 | 1489 | 0.61 | 2150 | 0.89 |  |
| DCSI |  |  |  |  |  |  |  |  |  |  |  |  |  |
| 1 | 68916 | 74.05 | 34535 | 58.89 | 30107 | 54.86 | 18960 | 52.77 | 188871 | 77.88 | 152518 | 62.89 | <0.001 |
| 2 | 12910 | 13.87 | 11329 | 19.32 | 11033 | 20.10 | 7141 | 19.88 | 28234 | 11.64 | 42413 | 17.49 |  |
| ≥3 | 11236 | 12.07 | 12777 | 21.79 | 13742 | 25.04 | 9828 | 27.35 | 25409 | 10.48 | 47583 | 19.62 |  |
| Medication |  |  |  |  |  |  |  |  |  |  |  |  |  |
| Metformin | 11329 | 12.17 | 5385 | 9.18 | 5101 | 9.29 | 4547 | 12.66 | - | - | 26362 | 10.87 | <0.001 |
| Sulfonamides | 16117 | 17.32 | 5821 | 9.93 | 3532 | 6.44 | 2054 | 5.72 | - | - | 27524 | 11.35 | <0.001 |
| TZD | 811 | 0.87 | 593 | 1.01 | 397 | 0.72 | 232 | 0.65 | - | - | 2033 | 0.84 | <0.001 |
| DPP-4i | - | - | 10 | 0.02 | 484 | 0.88 | 951 | 2.65 | - | - | 1445 | 0.60 | <0.001 |
| AGI | 751 | 0.81 | 661 | 1.13 | 567 | 1.03 | 400 | 1.11 | - | - | 2379 | 0.98 | <0.001 |
| SGLT-2 inhibitors | - | - | - | - | - | - | 65 | 0.18 | - | - | 65 | 0.03 | <0.001 |
| Number of oral antidiabetic drugs |  |  |  |  |  |  |  |  |  |  |  |  |  |
| 1 | 83505 | 89.73 | 54928 | 93.67 | 52115 | 94.96 | 33804 | 94.09 | - | - | 224352 | 92.51 | <0.001 |
| 2 | 8561 | 9.20 | 3097 | 5.28 | 2162 | 3.94 | 1542 | 4.29 | - | - | 15362 | 6.33 |  |
| ≥3 | 996 | 1.07 | 616 | 1.05 | 605 | 1.10 | 583 | 1.62 | - | - | 2800 | 1.15 |  |
| GLP1-RA | - | - | - | - | - | - | 8 | 0.02 | - | - | 8 | 0.00 | 0.0047 |
| Insulin | 13903 | 14.94 | 15926 | 27.16 | 15971 | 29.10 | 10550 | 29.36 | - | - | 56350 | 23.24 | <0.001 |
| ACEI/ARB | 17915 | 19.25 | 19605 | 33.43 | 20981 | 38.23 | 15144 | 42.15 | 30236 | 12.47 | 73645 | 30.37 | <0.001 |
| β-blockers | 12871 | 13.83 | 16595 | 28.30 | 17474 | 31.84 | 11452 | 31.87 | 31768 | 13.10 | 58392 | 24.08 | <0.001 |
| Calcium-channel<br>blockers | 23812 | 25.59 | 25722 | 43.86 | 26358 | 48.03 | 17529 | 48.79 | 43723 | 18.03 | 93421 | 38.52 | <0.001 |
| Diuretics | 15990 | 17.18 | 19146 | 32.65 | 18681 | 34.04 | 12005 | 33.41 | 34037 | 14.04 | 65822 | 27.14 | <0.001 |
| Number of antihypertensive drugs |  |  |  |  |  |  |  |  |  |  |  |  |  |
| 1 | 72195 | 77.58 | 33916 | 57.84 | 29316 | 53.42 | 18536 | 51.59 | 204010 | 84.12 | 153963 | 63.49 | <0.001 |
| 2 | 11616 | 12.48 | 10418 | 17.77 | 10467 | 19.07 | 7304 | 20.33 | 21084 | 8.69 | 39805 | 16.41 |  |
| ≥3 | 9251 | 9.94 | 14307 | 24.40 | 15099 | 27.51 | 10089 | 28.08 | 17420 | 7.18 | 48746 | 20.10 |  |
| Statin | 5770 | 6.20 | 9889 | 16.86 | 12627 | 23.01 | 9668 | 26.91 | 13848 | 5.71 | 37954 | 15.65 | <0.001 |
| Aspirin | 16311 | 17.53 | 18208 | 31.05 | 17328 | 31.57 | 11146 | 31.02 | 31380 | 12.94 | 62993 | 25.97 | <0.001 |
| ADP receptor<br>inhibitors | 184 | 0.20 | 756 | 1.29 | 980 | 1.79 | 891 | 2.48 | 1145 | 0.47 | 2811 | 1.16 | <0.001 |
| Vitamin K antagonists | 415 | 0.45 | 516 | 0.88 | 540 | 0.98 | 397 | 1.10 | 1181 | 0.49 | 1868 | 0.77 | <0.001 |
| Direct oral | - | - | - | - | 20 | 0.04 | 154 | 0.43 | 94 | 0.04 | 174 | 0.07 | <0.001 |
| anticoagulants |  |  |  |  |  |  |  |  |  |  |  |  |  |

Data are shown as n (%) or mean ± SD. T2D, type 2 diabetes; COPD, chronic obstructive pulmonary disease; PAOD, peripheral arterial occlusion disease; CCI, Charlson Comorbidity Index; DCSI, Diabetes Complications Severity Index; SGLT-2, sodium glucose cotransporter-2, GLP-1 RA, glucagon-like peptide-1 receptor agonist; ACEI, angiotensin-converting enzyme inhibitor; ARB, angiotensin receptor blocker. A p-value of >0.05 indicates a negligible difference between patients with T2D and non-T2D.

### Main Outcomes

We examined the incidence of in-hospital ischemic stroke and 30-day mortality after stroke admission from 2000 to 2019. Previous studies conducted in Taiwan have shown the reliability and accuracy of ICD codes in identifying ischemic stroke cases.^14,17^ Mortality data were confirmed using the National Death Registry. Incidence rates of ischemic stroke and associated 30-day mortality were calculated and compared between the study and control groups. Each participant was followed until the occurrence of one of the prespecified outcomes, withdrawal from the NHI system, or the end of the study on December 31, 2019, whichever occurred first.

### Statistical Analyses

We used 1:1 propensity score matching to adjust for year of recruitment, age, and sex between patients with and without T2D to improve comparability. The nearest neighbor technique was used for creating matched pairs, assuming that a p-value > 0.05 indicated balanced distributions in the study and control groups. For descriptive statistics, chi-squared tests and Student’s t-tests were used to analyze the distribution of categorical and continuous variables, respectively. Crude and multivariate-adjusted Cox proportional hazards models, accompanied by robust sandwich standard error estimates, were used to assess the differential risk of outcomes between the study and control groups. Verification of the proportional hazards assumption was facilitated by Schoenfeld residuals and complementary log-log plots.

Results were expressed as hazard ratios (HRs) and 95% confidence intervals (CIs) for the study and control groups.

We used Cox proportional hazards models to analyze differences in the risk of ischemic stroke and 30-day mortality between men and women and between patients aged <60 and those aged ≥60. We calculated the absolute and percent change in outcomes, the p-linear trend to see trend changes among these recruitment years, and the relative risk of outcomes between patients with and without T2D. We plotted figures to describe the relative risk of outcomes between patients (overall, male and female, age <60 and ≧60 years) with and without T2D among these recruiting years. We also performed a subgroup analysis to compare the risk of in-hospital ischemic stroke in patients under 60 years with T2D versus no T2D. A two-tailed p-value less than .05 was deemed statistically significant. All analyses were performed using SAS version 9.4 (SAS Institute Inc., Cary, NC, USA).

## RESULTS

### Baseline Characteristics of the Study Population

From the National Health Insurance Research Database for the years 2000-2004, 2005-2009, 2010-2014, and 2015-2018, we individually included 93,062, 58,641, 54,882, and 35,929 patients, respectively. Using propensity score matching for sex, age, and year of enrollment, we identified 242,514 pairs of patients with and without type 2 diabetes. Both groups showed excellent matching for age and sex (Table 1, p>0.05). In the matched cohort, 45.52% were female, the mean age was 54.23 years, and the mean follow-up for this study was 9.2 years.

### Trend of Ischemic Stroke

Between 2000 and 2018, the incidence of in-hospital ischemic stroke in patients with and without T2D had significant reductions of 64.42% and 55.63%, respectively. However, the relative risk of stroke associated with diabetes remained significantly higher (2.01 to 2.33 times) than that for non-diabetes (Table 2, Figure 1). Men had a higher risk of ischemic stroke than women. The relative risk of stroke was the same for patients with T2D (male or female) and those without T2D. However, from 2015 to 2018, there was a slight increase in the relative risk of stroke (relative risk = 2.86) for women with T2D compared to those without T2D.

**Figure 1.**
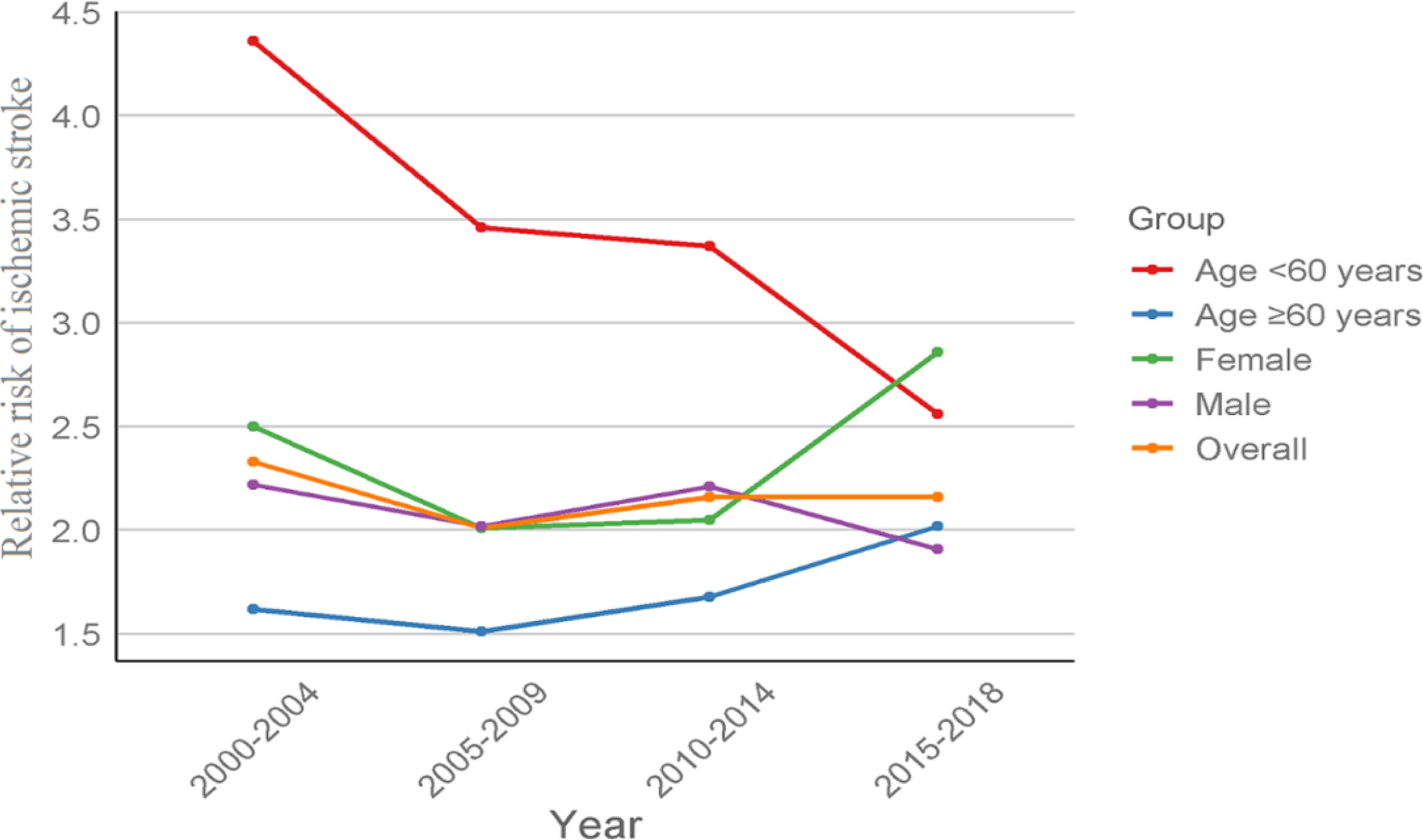
Relative risk of in-hospital ischemic stroke in patients with and without T2D in Taiwan from 2000-2018.

**Table 2.**
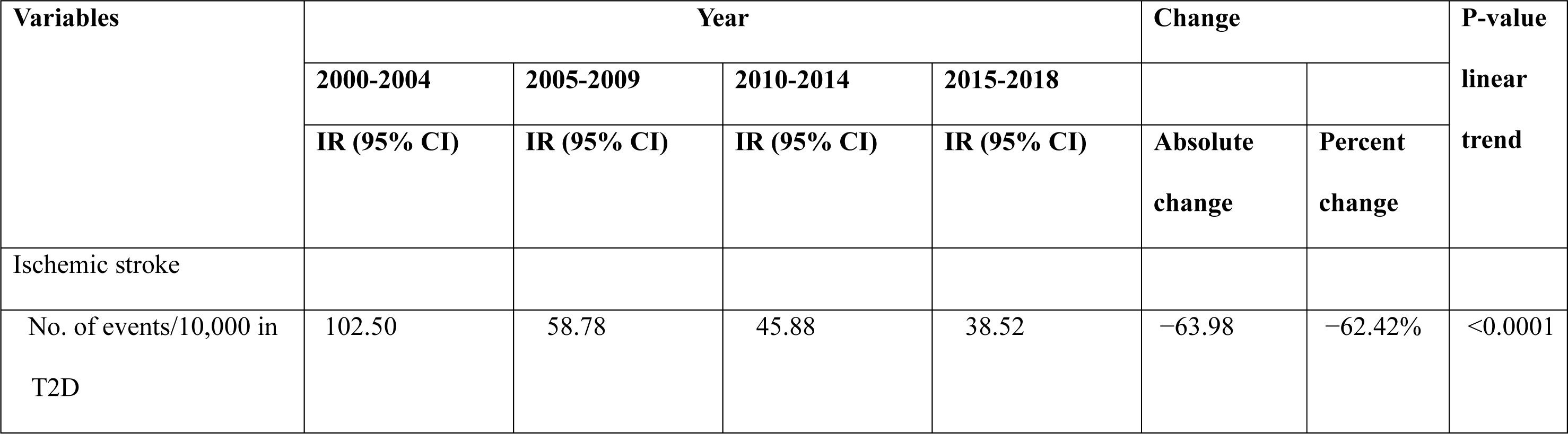
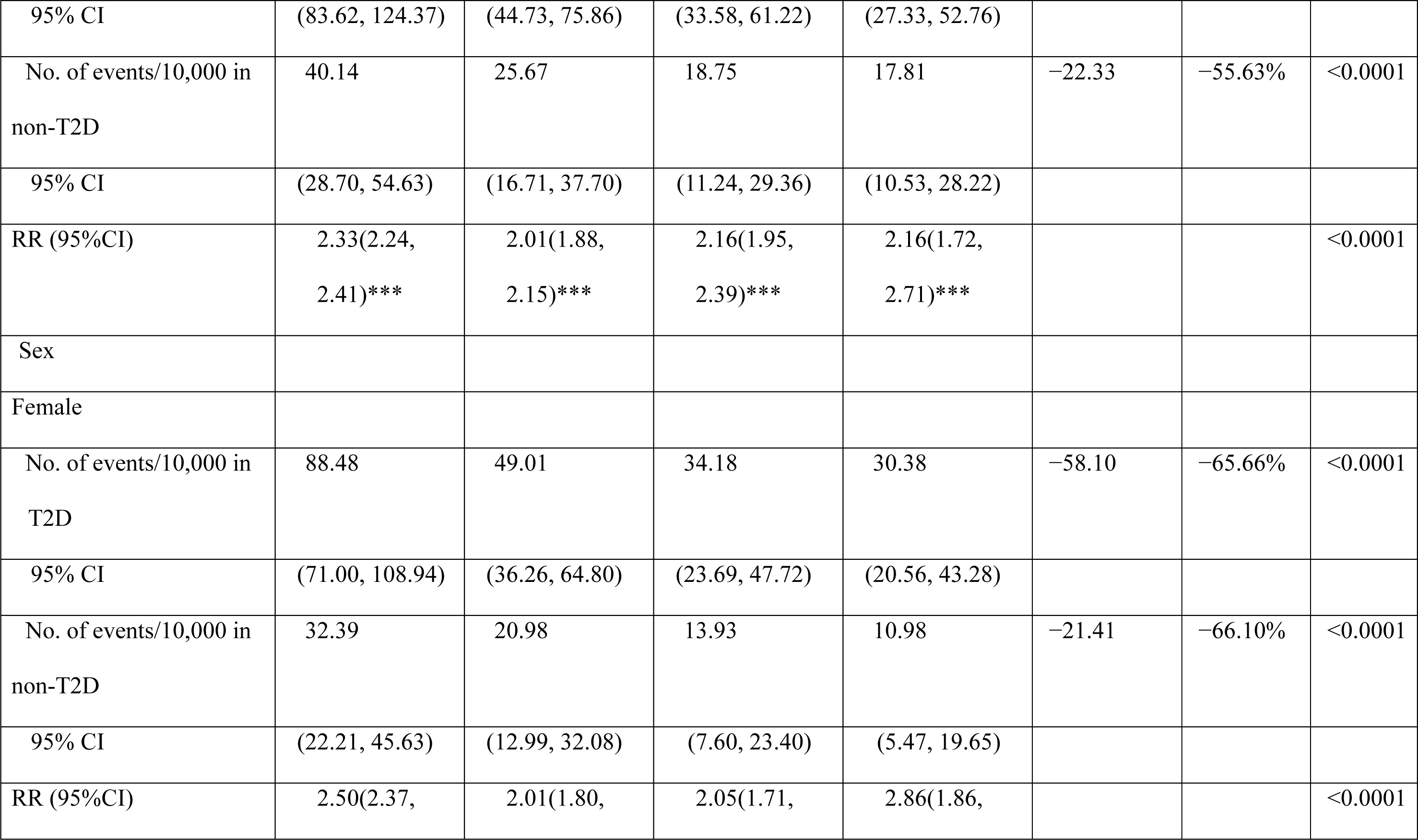
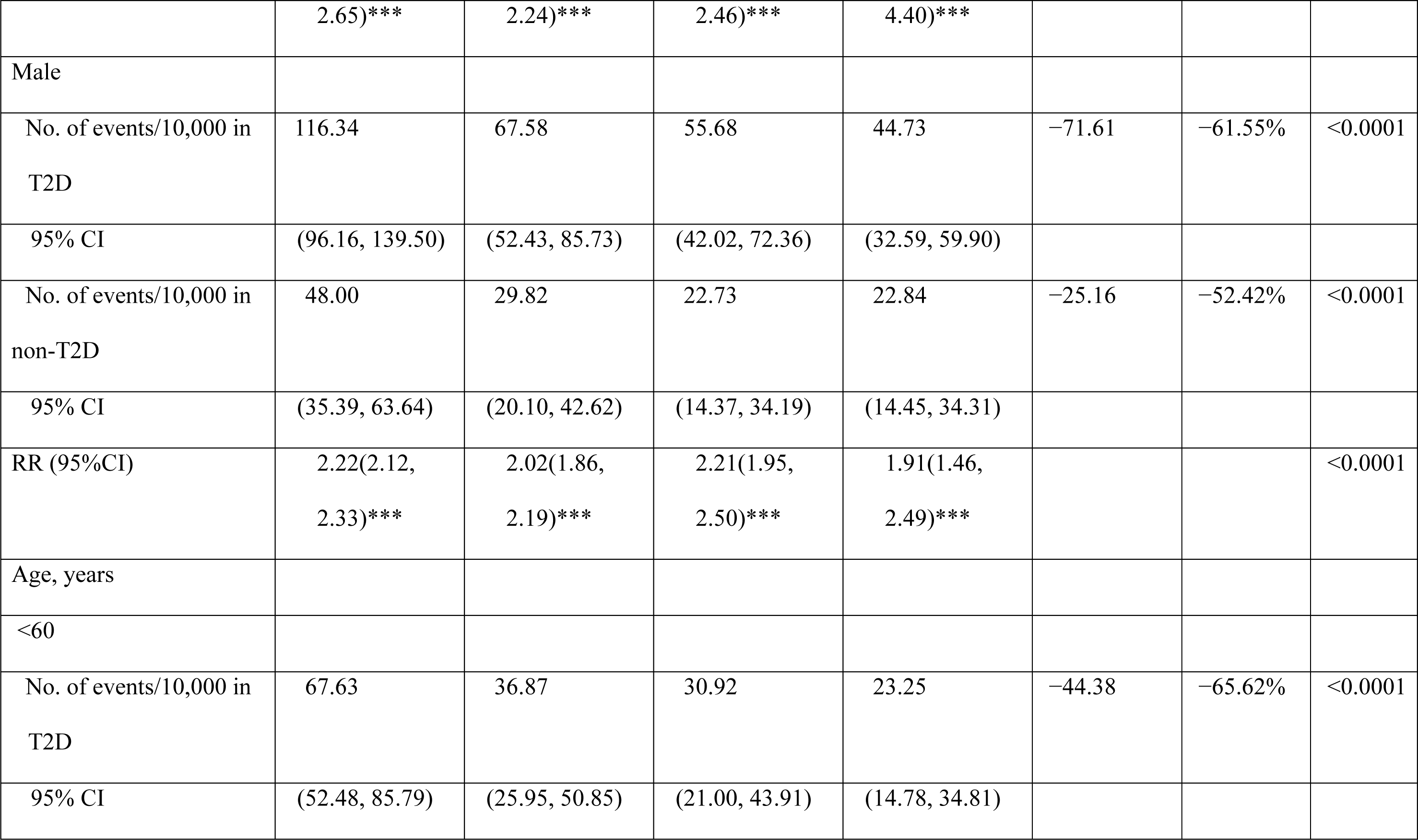
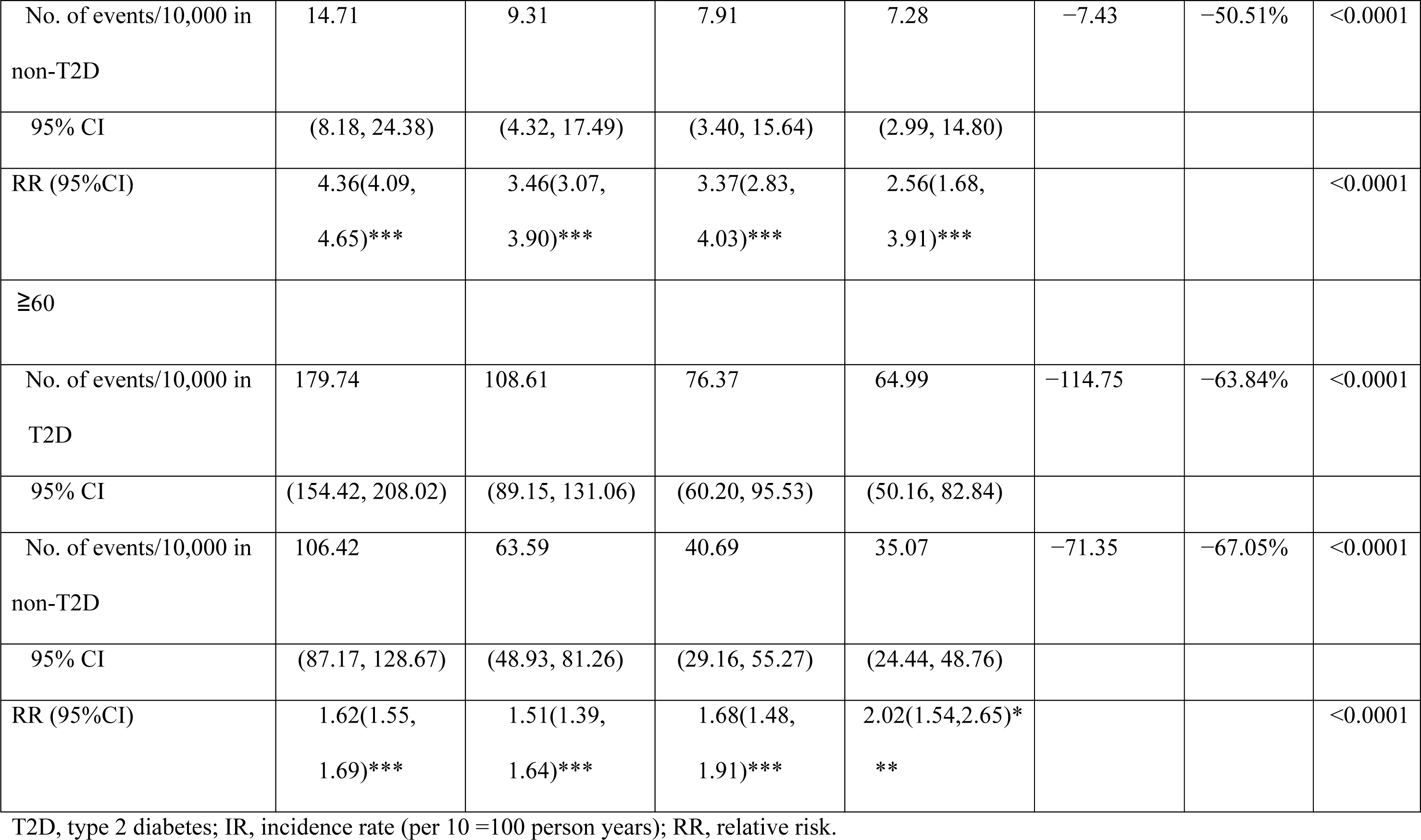
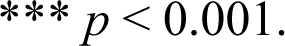
Event Rate and Relative Risk of Ischemic Stroke in Persons with and Without Type 2 Diabetes.

People with diabetes aged 60 years and older have a 1.51 to 2.02 times higher relative risk of stroke than people without diabetes. Conversely, people under 60 with diabetes have a more profound (ranging from 2.56 to 4.36 times) higher relative risk of stroke than those without diabetes. Nevertheless, between 2000 and 2018, the incidence of stroke associated with diabetes and non-diabetes in people under 60 decreased by 65.62% and 50.51%, respectively, resulting in a significant downward trend in the relative risk of stroke for people under 60 with diabetes compared to those without diabetes (Figure 1).

Among patients under 60, patients with diabetes between 18-39 years have a significantly higher relative risk of ischemic stroke than non-diabetics than those in the 40-59 age group. Female with diabetes have a significantly higher relative risk of ischemic stroke compared to non-diabetics than their male counterparts (Table S2).

### 30-Day Mortality Rate

Between 2000 and 2018, there was a significant decrease in the 30-day mortality rate after stroke in patients with and without T2D, with a reduction of 83.24% and 88.55%, respectively, and no significantly higher risk of death from stroke in people with diabetes compared to those without diabetes. The risk of death from stroke was similar for men and women. However, there was an abrupt increase in the relative risk of death for women with T2D compared to women without T2D from 2015 to 2018. Among those aged 60 and older, patients with diabetes had the same or even lower relative risk of death from stroke compared to non-diabetics. However, those under 60 with diabetes had a 1.63 to 2.49 times higher relative risk of dying from stroke than their non-diabetic counterparts (Table 3, Figure 2).

**Figure 2.**
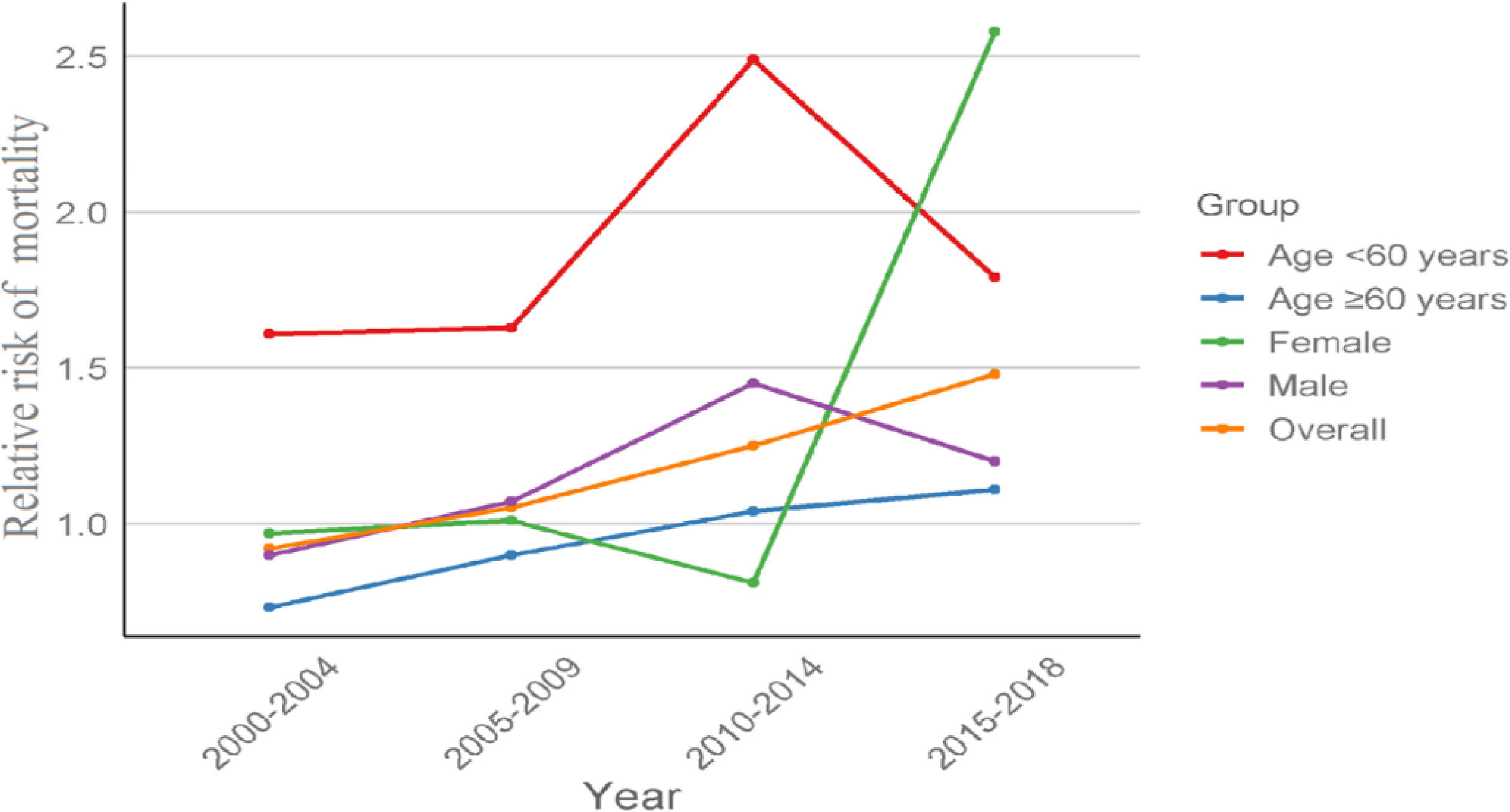
**Relative risk of 30-day mortality after stroke in patients with and without T2D in Taiwan from 2000-2018.**

**Table 3.** Event Rate and Relative Risk of 30-Day Mortality After Ischemic Stroke in Persons with and Without Type 2 Diabetes.

| Variables | Year |  |  |  | Change |  | P-value<br>linear<br>trend |
| --- | --- | --- | --- | --- | --- | --- | --- |
|  | 2000-2004 | 2005-2009 | 2010-2014 | 2015-2018 |  |  |  |
|  | IR (95% CI) | IR (95% CI) | IR (95% CI) | IR (95% CI) | Absolute<br>change | Percent<br>change |  |
| 30-day mortality |  |  |  |  |  |  |  |
| No. of events/10,000 in T2D | 4.73 | 2.56 | 1.70 | 0.79 | −3.94 | −83.24% | <0.0001 |
| 95% CI | (1.47, 11.28) | (0.44, 8.10) | (0.16, 6.74) | (0.01, 5.21) |  |  |  |
| No. of events/10,000 in<br>non-T2D | 4.50 | 2.21 | 1.24 | 0.52 | −3.98 | −88.55% | <0.0001 |
| 95% CI | (1.35, 10.97) | (0.31, 7.55) | (0.06, 5.98) | (0.00, 4.70) |  |  |  |
| RR (95%CI) | 0.92(0.81, 1.05) | 1.05(0.81,<br>1.38) | 1.25(0.80,<br>1.96) | 1.48(0.35,<br>6.25) |  |  | <0.0001 |
| Sex |  |  |  |  |  |  |  |
| Female |  |  |  |  |  |  |  |
| No. of events/10,000 in T2D | 4.65 | 2.37 | 1.10 | 0.61 | -4.04 | -86.85% | <0.0001 |
| 95% CI | (1.43, 11.17) | (0.37, 7.81) | (0.04, 5.74) | (0.00, 4.88) |  |  |  |
| No. of events/10,000 in non-T2D | 4.11 | 2.06 | 1.02 | 0.30 | -3.81 | -92.59% | <0.0001 |
| 95% CI | (1.15, 10.40) | (0.26, 7.32) | (0.03, 5.60) | (0.00, 4.30) |  |  |  |
| RR (95%CI) | 0.97(0.80, 1.17) | 1.01(0.67, 1.53) | 0.81(0.35, 1.90) | 2.58(0.19, 35.64) |  |  | <0.0001 |
| Male |  |  |  |  |  |  |  |
| No. of events/10,000 in T2D | 4.81 | 2.73 | 2.20 | 0.93 | -3.88 | -80.63% | <0.0001 |
| 95% CI | (1.52, 11.40) | (0.51, 8.36) | (0.31, 7.54) | (0.02, 5.45) |  |  |  |
| No. of events/10,000 in non-T2D | 4.90 | 2.34 | 1.42 | 0.67 | -4.23 | -86.33% | <0.0001 |
| 95% CI | (1.57, 11.53) | (0.36, 7.75) | (0.09, 6.28) | (0.00, 4.99) |  |  |  |
| RR (95%CI) | 0.90(0.75, 1.07) | 1.07(0.76, 1.52) | 1.45(0.85, 2.47) | 1.20(0.21, 6.96) |  |  | <0.0001 |
| Age, years |  |  |  |  |  |  |  |
| <60 |  |  |  |  |  |  |  |
| No. of events/10,000 in T2D | 2.35 | 1.01 | 0.66 | 0.63 | -1.72 | -73.34% | <0.0001 |
| 95% CI | (0.36, 7.77) | (0.03, 5.59) | (0.00, 4.97) | (0.00, 4.91) |  |  |  |
| No. of events/10,000 in non-diabetes | 1.40 | 0.60 | 0.26 | 0.62 | -0.78 | -55.60% | <0.0001 |
| 95% CI | (0.09, 6.26) | (0.00, 4.86) | (0.00, 4.21) | (0.00, 4.90) |  |  |  |
| RR (95%CI) | 1.61(1.27, 2.06)*** | 1.63(0.94, 2.80) | 2.49(0.88, 7.03) | 1.79(1.15, 3.50) |  |  | <0.0001 |
| ≥60 |  |  |  |  |  |  |  |
| No. of events/10,000 in T2D | 9.77 | 5.99 | 3.80 | 1.08 | -8.69 | -88.94% | <0.0001 |
| 95% CI | (4.64, 18.09) | (2.20, 13.04) | (0.99, 9.95) | (0.03, 5.71) |  |  |  |
| No. of events/10,000 in | 12.34 | 5.85 | 3.21 | 1.36 | -10.98 | -89.01% | <0.0001 |
| non-T2D |  |  |  |  |  |  |  |
| 95% CI | (6.45, 21.40) | (2.11, 12.85) | (0.71, 9.08) | (0.08, 6.18) |  |  |  |
| RR (95%CI) | 0.73(0.62,<br>0.85)*** | 0.90(0.66,<br>1.23) | 1.04(0.63,<br>1.72) | 1.11(0.23,<br>5.40) |  |  | <0.0001 |

T2D, type 2 diabetes; IR, incidence rate; RR, relative risk.
\*\*\* $p < 0.001$ .

## DISCUSSION

From 2000 to 2018, the incidence rate of in-hospital ischemic stroke has shown a significantly decreasing trend in patients with T2D and those without T2D in Taiwan. However, people with T2D still have a significantly higher risk of stroke than those without T2D, which is particularly notable for people under 60 with T2D, who have a higher risk of stroke than those without T2D. Patients with and without T2D in Taiwan experienced a significant decrease in 30-day mortality after ischemic stroke from 2000 to 2018. Patients under 60 with T2D have a higher risk of dying within 30 days of stroke than those without T2D. However, people over 60 with T2D have a similar or slightly lower risk of 30-day mortality than those without T2D.

In recent decades, the lifetime risk of stroke has declined in some high-income countries.

However, the global lifetime risk of stroke continues to rise, possibly due to an aging population and increased incidence of metabolic diseases.^4^ People with diabetes have twice the risk of ischemic stroke than those without diabetes.^5,6^ In many countries, the risk of ischemic stroke from diabetes has decreased.^7,18–23^ However, it remains about the same in some countries,^9^ and in others, it has increased.^11^ The varying trends in the incidence of ischemic stroke in different countries may be related to different definitions of diabetes and stroke, the control of risk factors, and differences in environmental and genetic factors.^21^ In Taiwan, we observed a notable decline in the incidence of ischemic stroke from 2000 to 2018 in individuals with T2D and those without T2D. However, the decline was slightly more pronounced in individuals with diabetes than in those without diabetes. Nevertheless, people with diabetes still have more than twice the risk of having a stroke than people without diabetes. The decline in the incidence rate of ischemic stroke may be related to improved control of blood glucose, blood pressure, and cholesterol levels in recent years in Taiwan.^24,25^

Men have a higher risk of stroke than women in Taiwan. However, the relative risk of stroke in people with diabetes compared to those without diabetes and the declining trend is consistent for both men and women, in line with the overall population trend. This finding is consistent with the results of the Korea^20^ and Scotland^21^ studies. Because of younger age, people under 60 have a lower incidence of ischemic stroke than those over 60. However, people with diabetes under 60 have a significantly higher risk of stroke (ranging from 2.56 to 4.36 times) than those without diabetes. This finding is consistent with previous studies.^5,6,20,22^ However, from 2000 to 2018, the decline in stroke incidence rate increased in people with diabetes under 60 compared to those without diabetes. Therefore, in Taiwan, the relative risk of experiencing an ischemic stroke has gradually decreased among individuals under 60 with diabetes compared to those without the disease.

Diabetes can worsen the prognosis for patients with stroke by increasing the likelihood of disability and even increasing the mortality rate from stroke.^5^ In Taiwan, from 2000 to 2018, there has been a significant improvement in the 30-day mortality rate for stroke in individuals with and without diabetes, probably due to the implementation of the acute stroke care program in Taiwan in recent years, which includes the use of thrombolytic treatments, improved management of severe strokes, and higher identification of minor strokes.^26^ Diabetes does not appear to have a higher stroke mortality rate than non-diabetes, consistent with the results of some previous studies.^27–29^ The 30-day mortality rate is similar for men and women. However, between 2015 and 2018, there was a sudden increase in the relative risk of stroke mortality in women with diabetes compared to women without diabetes. The reason for this is unclear. Individuals over 60 with diabetes have the same or a lower relative risk of stroke mortality compared to those without diabetes. However, people under 60 with diabetes still have a 1.63 to 2.49 times higher relative risk of stroke mortality than those without diabetes. We should work to improve the management of stroke in people under 60 with diabetes and women to reduce mortality.

Our study has some strengths and clinical implications. First, the Taiwan National Health Insurance Research Database covers the medical records of 99% of the population, which can help reduce selection bias. The study uses a nationwide database, an excellent tool for monitoring national trends in stroke incidence and related mortality. As this database is linked to the National Death Registry, the observations on the risk of stroke mortality are relatively reliable. Second, we observe that individuals under 60 with T2D have a significantly higher relative risk of ischemic stroke and a higher 30-day mortality rate than those without T2D. Therefore, we should encourage people under 60 with diabetes to increase physical activity levels, reduce obesity, stop smoking, avoid excessive alcohol consumption, and maintain optimal blood glucose, blood pressure, and cholesterol levels.

Using medications such as pioglitazone or glucagon-like peptide-1 receptor agonists (GLP-1 RA) to manage diabetes may be a proactive approach to prevent stroke development and improve prognosis.^30,31^

This study has several limitations. First, the dataset does not include information on blood pressure, glucose, hemoglobin A1C, biochemical results, medical images, and echocardiograms, which are essential for proper diagnosis and assessment of T2D, hypertension, and stroke. Nevertheless, we used ICD codes to identify these conditions, a method validated with good accuracy in previous studies.^14,17^ The lack of details on blood pressure and glucose levels implies that we could not assess treatment progress or measure the severity of hypertension and diabetes in the study. However, to reduce the potential confounding effects of hypertension and T2D, we adjusted for the type and number of antihypertensive and antidiabetic medications used in our multivariable analyses. Second, our administrative dataset does not include details of family history, physical activity, and eating habits. Nevertheless, we included important factors such as age, sex, comorbidities, and use of antidiabetic, antihypertensive, and anticoagulant medications to adjust for multivariable analyses when comparing the study and control groups. Third, the participants in this study are predominantly Taiwanese; therefore, the results may not directly apply to other ethnic groups. However, they serve as a valuable source of information on stroke data in the Eastern population. Finally, this cohort study inherently contains some unknown confounders.

Therefore, the results can only indicate associations and not establish causal relationships.

## CONCLUSIONS

This nationwide population-based cohort study showed that from 2000 to 2018, the incidence rate of ischemic stroke exhibited a significant decreasing trend in patients with T2D and those without T2D in Taiwan. However, patients with T2D still have a significantly higher risk of ischemic stroke than those without T2D, which is particularly notable for patients under 60 with T2D, who have a higher risk of stroke than those without T2D. Patients with and without T2D in Taiwan showed a significant decrease in the 30-day mortality rate after stroke from 2000 to 2018. Patients younger than 60 with T2D have a higher risk of 30-day mortality than those without T2D. We should encourage patients under 60 with T2D to adopt a healthy lifestyle and optimize the management of diabetes, hypertension, and dyslipidemia to reduce the risk of ischemic stroke and mortality.

## Abbreviations

COPD: chronic obstructive pulmonary disease
CCI: Charlson comorbidity Index
DCSI: diabetes complications severity index
GLP-1: RAglucagon-like peptide 1 receptor agonist;
T2D: type 2 diabetes.

## Disclosure

The authors declare no competing interests.

## Data Availability

Data of this study are available from the National Health Insurance Research Database (NHIRD) published by Taiwan National Health Insurance (NHI) Administration. The data utilized in this study cannot be made available in the paper, the supplemental files, or in a public repository due to the ??Personal Information Protection Act?? executed by Taiwan government starting from 2012. Requests for data can be sent as a formal proposal to the NHIRD Office (https://dep.mohw.gov.tw/DOS/cp-2516-3591-113.html) or by email to.

## Acknowledgments

We are grateful to Health Data Science Center, China Medical University Hospital, Szu-Yuan Research Foundation of Internal Medicine, and Y.L. Lin Hung Tai Education Foundation for providing administrative, technical and funding support. The funders had no role in the study design, data collection and analysis, the decision to publish, or preparation of the manuscript. No additional external funding was received for this study.

## Supplemental Material

Tables S1–S2

